# Effectiveness of isolation, testing, contact tracing and physical distancing on reducing transmission of SARS-CoV-2 in different settings

**DOI:** 10.1101/2020.04.23.20077024

**Authors:** Adam Kucharski, Petra Klepac, Andrew Conlan, Stephen Kissler, Maria Tang, Hannah Fry, Julia Gog, John Edmunds, CMMID COVID-19 working group

**Affiliations:** Centre for Mathematical Modelling of Infectious Diseases, London School of Hygiene & Tropical Medicine; Department of Veterinary Medicine, University of Cambridge; Department of Immunology and Infectious Diseases, Harvard T.H. Chan School of Public Health; Department of Applied Mathematics and Theoretical Physics, University of Cambridge; Centre for Advanced Spatial Analysis, University College London

## Abstract

**Background:** Isolation of symptomatic cases and tracing of contacts has been used as an early COVID-19 containment measure in many countries, with additional physical distancing measures also introduced as outbreaks have grown. To maintain control of infection while also reducing disruption to populations, there is a need to understand what combination of measures – including novel digital tracing approaches and less intensive physical distancing – may be required to reduce transmission.

**Methods:** Using a model of individual-level transmission stratified by setting (household, work, school, other) based on BBC Pandemic data from 40,162 UK participants, we simulated the impact of a range of different testing, isolation, tracing and physical distancing scenarios. As well as estimating reduction in effective reproduction number, we estimated, for a given level of COVID-19 incidence, the number of contacts that would be newly quarantined each day under different strategies.

**Results:** Under optimistic but plausible assumptions, we estimated that combined testing and tracing strategies would reduce transmission more than mass testing or self-isolation alone (50–65% compared to 2–30%). If limits are placed on gatherings outside of home/school/work (e.g. maximum of 4 daily contacts in other settings), then manual contact tracing of acquaintances only could have a similar effect on transmission reduction as detailed contact tracing. In a scenario where there were 10,000 new symptomatic cases per day, we estimated in most contact tracing strategies, 140,000 to 390,000 contacts would be newly quarantined each day.

**Conclusions:** Consistent with previous modelling studies and country-specific COVID-19 responses to date, our analysis estimates that a high proportion of cases would need to self-isolate and a high proportion of their contacts to be successfully traced to ensure an effective reproduction number that is below one in the absence of other measures. If combined with moderate physical distancing measures, self-isolation and contact tracing would be more likely to achieve control.

**Funding:** Wellcome Trust, EPSRC, European Commission.

## Introduction

The novel SARS-CoV-2 coronavirus spread rapidly across multiple countries in early 2020 (1–3). A staple public health control measure for outbreaks of emerging directly-transmitted infections involves isolation of symptomatic cases as well as tracing, testing and quarantine of their contacts (2). The effectiveness of this measure in containing new outbreaks depends both on the transmission dynamics of the infection and the proportion of transmission that occurs from infections without symptoms (4). There is evidence that SARS-CoV-2 has a reproduction number of around 2–3 in the early stages of an outbreak (1,5) and many infections can occur without symptoms (6), which means isolation of symptomatic cases and contact tracing alone are unlikely to contain an outbreak unless a high proportion of cases are isolated and contacts successfully traced and quarantined (7).

Several countries have used combinations of non-pharmaceutical interventions to reduce SARS-CoV-2 transmission (3,8–11). As well as isolation of symptomatic individuals and tracing and quarantine of their contacts, measures have included general physical distancing, school closures, remote working, community testing and cancellation of events. It has also been suggested that the effectiveness of contact tracing could be enhanced through app-based digital tracing (12,13). The effectiveness of contact tracing and the extent of resources required to implement it successfully will depend on the social interactions within a population (14). Targeted interventions such as contact tracing also need to consider individual-level variation in transmission: high variation can lead to superspreading events, which could result in larger numbers of contacts needing to be traced (15,16). There are several examples of such events occurring for COVID-19, including meals, parties and other social gatherings involving close contacts (17–19).

We used social contact data from a large-scale UK study of over 40,000 participants (20,21) to explore a range of different control measures for SARS-CoV-2, including: self-isolation of symptomatic cases; household quarantine; manual tracing of acquaintances (i.e. contacts that have been met before); manual tracing of all contacts; app-based tracing; mass testing regardless of symptoms; a limit on daily contacts made outside home, school and work; and having proportion of the adult population work from home. As well as estimating the reduction in transmission under different scenarios, we estimated how many primary cases and contacts would be quarantined per day in different strategies for a given level of symptomatic case incidence.

## Research in context

### Evidence before this study

There is an extensive literature on isolation and contact tracing for pathogens such as SARS, smallpox and Ebola. Early modelling studies of SARS-CoV-2 suggested that isolation and tracing alone may not be sufficient to control outbreaks, and additional measures may be required; these measures have since been explored in population-level models. However, there has not been an analysis using setting-specific social contact data to quantify the potential impact of combined contact tracing and physical distancing measures on reducing individual-level transmission of SARS-CoV-2.

### Added value of this study

We use data from over 40,000 individuals to assess contact patterns and SARS-CoV-2 transmission in different settings, and compare how combinations of self-isolation, contact tracing and physical distancing could reduce secondary cases. We assessed a range of combined physical distancing and testing/tracing measures, including app-based tracing, remote working, limits on different sized gatherings, and mass population-based testing. We also estimated the number of contacts that would be quarantined under different strategies.

### Implications of all the available evidence

Several characteristics of SARS-CoV-2 make effective isolation and contact tracing challenging, including high transmissibility, a relatively short serial interval, and transmission that can occur without symptoms. Combining isolation and contact tracing with physical distancing measures – particularly measures that reduce contacts in settings that would otherwise be difficult to trace – could therefore increase the likelihood of achieving sustained control.

## Methods

### Secondary attack rate data sources

To estimate the risk of transmission per contact in different community settings, we collated contact tracing studies for COVID-19 from multiple settings that stratified contacts within and outside households (Table 1). Across studies, the estimated secondary attack rate (SAR) within households was 10–20%, with a much smaller SAR among close contacts made outside households, with estimates of 0–5% across studies. However, all these studies were conducted in an ‘under control’ scenario (i.e. effective reproduction number R<1) and some reported relatively few contacts, which may omit superspreading events. This suggests that SARS-CoV-2 may be driven by community transmission events as well as household contacts (18). In our main analysis, we therefore assume 20% HH SAR and 6% among all contacts, which reflects the upper bound of estimates in observed studies (Table 1) and led to an overall reproduction number of 2.6 in our model (described in next section) when no control measures were in place.

**Table 1:**
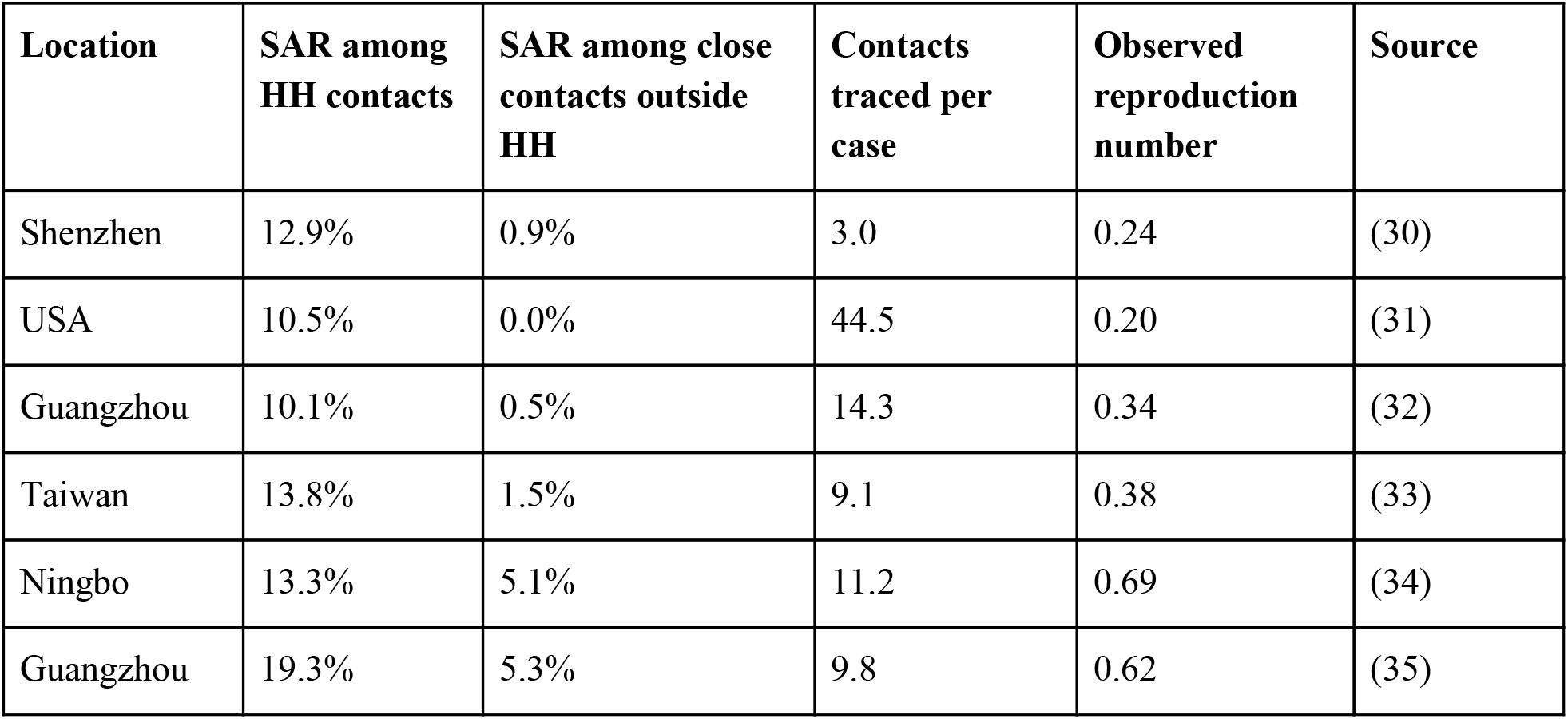
Secondary attack rates estimated from COVID-19 contact tracing studies.

### Transmission model

Our analysis is based on data on 40,162 UK participants with recorded social contacts in the BBC Pandemic dataset (20). Using these data, we simulated a large number of individual-level transmission events by repeatedly generating contact distributions for a primary case and randomly generating infections among these contacts. In each simulation, we randomly specify a primary case as either under 18 or 18 and over, based on UK demography, in which 21% of the population are under 18 (22). We then generate contacts by randomly sampling values from the marginal distributions of daily contacts made in three different settings for their age group (i.e. under 18 or adults): at home; at work & school; and in ‘other’ settings (Figure 1A–B). We used the marginal distributions rather than raw participant data to ensure non-identifiability and reproducibility in our model code.

**Figure 1:**
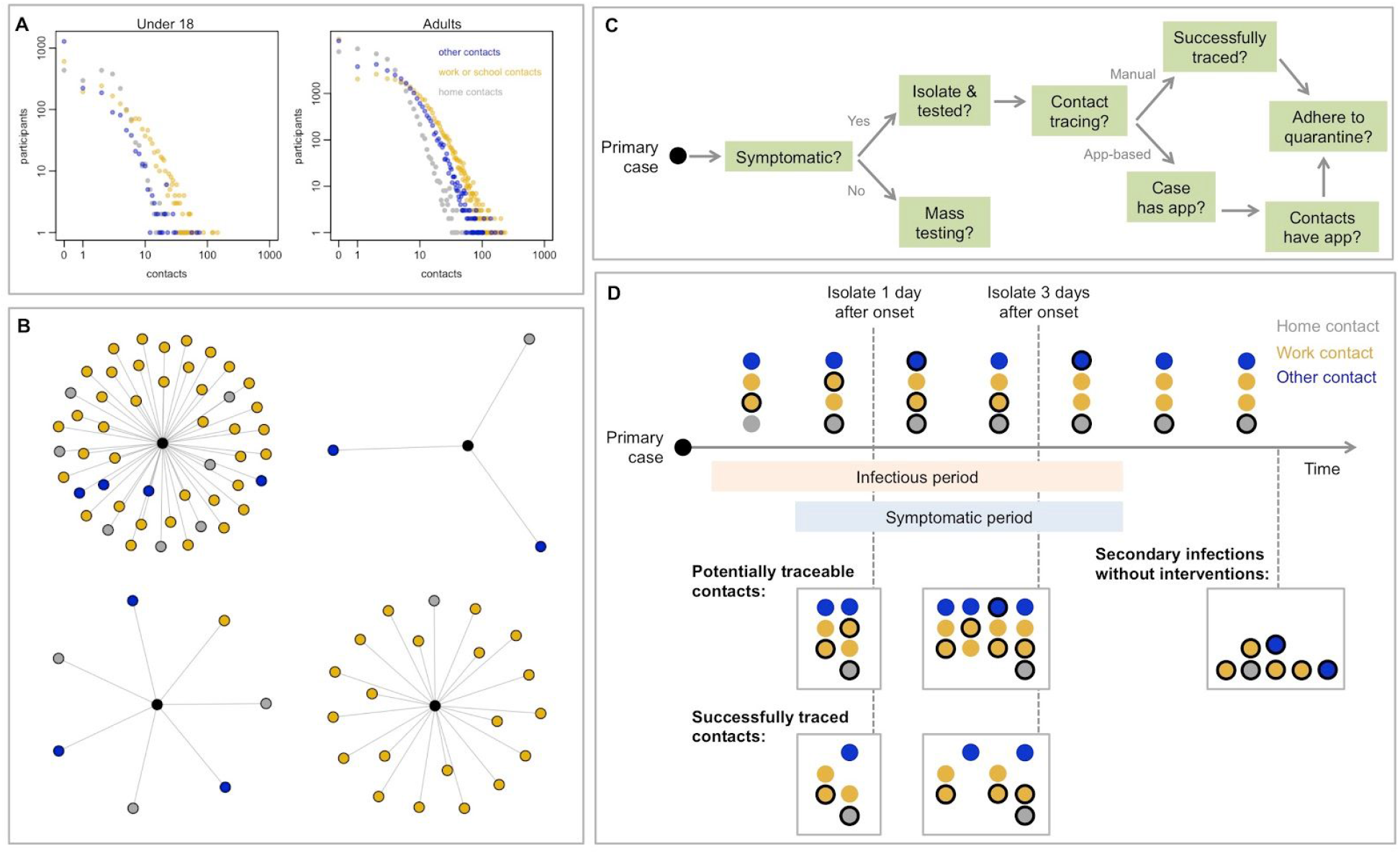
Model of social interactions and SARS-CoV-2 transmission and control. A) Distribution of daily contacts made at home, work/school and other settings in the BBC Pandemic dataset. B) Examples of daily social contact patterns for four randomly selected individuals in the model. Black point shows the individual reporting contacts, with social contacts coloured as in A. C) Factors that influence whether an individual is isolated and whether contacts are successfully traced in the model (parameters in Table 2). D) Implementation of contact tracing in the model. Timeline shows a primary case with four daily contacts self-isolating either 1 or 3 days after onset of symptoms. We assume the household contact is the same person throughout, whereas other contacts are made independently. Had the primary case not been isolated, there would have been 7 secondary cases in this illustration (shown with circulations). For isolation 1 day after onset, 4 secondary infections are prevented immediately. Then 7 contacts are potentially traceable, 3 of whom are infected. In this example, two infected contacts pre-isolation are successfully traced and quarantined (i.e. one is missed), so overall the isolation-and-tracing control measure results in a 4+2 = 6 reduction in effective reproduction number. A similar illustration is shown for isolation 3 days after onset.

In the model, we assumed infected individuals had a certain probability of being symptomatic and of being tested if symptomatic, as well as an infectious period that depended on when/if they self-isolated following onset of symptoms (details and justification for model parameters provided in Table 2). We assumed a mean delay of 2.6 days from onset-to-isolation in our baseline scenario (Figure S1). We assumed individuals became infectious one day before onset of symptoms. During each day of the effective infectious period, individuals made a given number of contacts equal to their simulated daily contacts. To avoid double-counting household members, contacts made within the home were not tallied over the entire infectious period, but instead were fixed at the daily value. Once the individual-level contacts had been defined, we generated secondary infections at random based on assumed secondary attack rates among contacts made in different settings, and estimated how many contacts would be successfully traced in each of these settings under different scenarios. First, we generated the number of secondary cases without any control measures in place. Second, we randomly sampled the proportion of these secondary cases that were either successfully traced and quarantined, and hence removed from the potentially infectious pool, or averted through isolation of the primary case. The difference between these two values gave the overall number of secondary cases that would contribute to further transmission (Figure 1C–D).

**Table 2:** Parameter definitions and assumptions for the baseline model.

| Parameter | Assumed value | Details & references |
| --- | --- | --- |
| ***Individual-level epidemiology*** |  |  |
| Reproduction number in absence of control measures | 2.6 | Consistent with meta-analysis of early studies (36). Sensitivity analysis shown in Table S1. |
| Duration of infectiousness | 5 days (for cases that will become symptomatic, 1st day is pre-symptomatic) | Given incubation period around 5 days (37), this assumption implies serial interval of around 6.5 days (30). Sensitivity analysis shown in Table S2. |
| Relative infectiousness of asymptomatic cases | 50% | Point estimate was 65% in (34), but secondary cases from asymptomatics were more likely to in turn be asymptomatic, suggesting lower contribution to transmission. Sensitivity analysis shown in Figure S2. |
| Proportion of cases that are eventually symptomatic | 60% | (3,38,39) |
| Probability symptomatic individual will eventually self-isolate and be tested | 90% | We assume virus is only detectable by PCR during the infectious period. 90% UK survey respondents said would likely comply with app request to self-isolate if rapid test available (40) |
| Effective duration of infectiousness if self-isolate when symptomatic | Mean delay from onset to isolation of 2.6 days. Distribution shown in Figure S1. | Assume most likely to self-isolate 0–4 days after onset (i.e. 1–5 days after becoming infectious). For 269 cases with known date of onset and confirmation in Singapore, of those who were confirmed within 5 days, 2% were confirmed on date of onset, 26% on second day, 27% on 3rd day, 14% on 4th day and 31% on 5th day (41). We assume isolation could occur 1 day before confirmation. |
| ***Contact tracing*** |  |  |
| Secondary attack rate among contacts in home | 20% | See ‘secondary attack rate’ section of methods. |
| Secondary attack rate among other contacts | 6% | See ‘secondary attack rate’ section of methods. |
| Proportion of contacts that are acquaintances (i.e. have been met before) | 100% in household<br>90% at school<br>79% at work<br>52% in other settings | In the BBC Pandemic dataset (20), 96% of contacts made in the home had been met before, but we assume 100% of those living in same household have met before. |
| Proportion of potentially traceable household contacts that are successfully traced | 100% | Assumed |
| Proportion of potentially traceable workplace, school or ‘other’ contacts that are successfully traced | 95% | Assumed, with sensitivity analysis shown in Figure 3. |
| Probability traced contacts adhere to quarantine | 90% | Proportion of traced contacts that are successfully removed from the potentially infectious group. Same justification as ‘Probability symptomatic individual will eventually self-isolate and be tested’ parameter above. |
| ***App-based tracing*** |  |  |
| Proportion of population that would have app | 53% (= 71% x 75%) | 85% of age 16+ in UK are smartphone users (Ofcom, 2019). 16% of UK are under 10 or over 80 (22), so we assume 71% of population use smartphones. 75% of UK survey respondents said would probably or definitely download app (40) |
| ***Mass testing*** |  |  |
| Proportion of population that are tested per week | 5% (i.e. 460,000 tests per day for UK) | 0.7% of population tested per day, i.e. equal to the highest number of daily per capita tests performed anywhere in world as of mid-April 2020 (Iceland, 7 per 1000) (26) |

In each simulation, for each of the four contact settings, the number of baseline secondary infections per primary case under no control measures were drawn from a binomial distribution *R*_*base*_ *= B(N*_*c*_, *p*_*inf*_), where *N*_*c*_ *= (number of daily contacts)* × *(days infectious)* and *p*_*inf*_ *= SAR* × *(relative infectiousness)*, where relative infectiousness *= 1* if an individual is (pre-)symptomatic and 50% if asymptomatic. We then generated secondary infections accounting for reduction in *R*_*isol*_ *= B(R*_*base*_, *1–p*_*isol*_ ), where *p*_*isol*_ is the proportion of the infectious period spent in isolation. In the household setting, we assume *N*_*c*_ *= (number of daily contacts)* because the household contacts will be repeated each day. The number of infected contacts successfully traced were in turn drawn from a binomial distribution *R*_*traced*_ *= B(R*_*isol*_, *p*_*trace*_), where *p*_*trace*_ *= P(successfully traced)* × *P(individual adheres full to quarantine)*. Hence the reduced effective individual-level secondary cases resulting from control measures was equal to *R*_*control*_ = *R*_*base*_ – *R*_*traced*_. The overall effective reproduction number *R*_*eff*_ under different control scenarios was equal to the mean of *R*_*control*_ across all simulations.

### Scenarios

We considered several different scenarios, both individually and in combination. These included: no control measures; self-isolation of symptomatic cases away from their household; self-isolation and household quarantine after onset of symptoms in primary case; quarantine of work/school contacts; manual tracing of acquaintances (i.e. contacts that have been met before); manual tracing of all contacts; app-based tracing; mass testing of cases regardless of symptoms; a limit on daily contacts made in ‘other’ settings (with the baseline limit being 4 contacts, equal to the mean number reported by adults in the BBC data); and a proportion of the adult population working from home. In the self-isolation only scenario, we assumed individuals who were successfully isolated had no risk of onward transmission (even to household members). Otherwise we assumed household quarantine was in place alongside other measures. For app-based tracing to be successfully implemented in a given simulation, both the infectious individual and their contacts needed to have and use the app. We assumed individuals under age 10 or over 80 would not use a smartphone app (Table 2). In the scenario with mass testing of cases regardless of symptoms, we assumed infected individuals would be identified and immediately self-isolate at a random point during or after their 5 day infectious period. We assumed that infected individuals would not test positive if tested during the latent period (defined here as 1 day prior to symptoms onset). No other measures (e.g. self-isolation/quarantine) were in place for this scenario. For each intervention scenario, we simulated 20,000 primary cases, generating individual-level contact distributions and secondary cases with and without the control measure in place, as described in the previous section. Model code is available from: https://github.com/adamkucharski/2020-cov-tracing

## Results

Under the control measures considered, we found that combined testing and tracing strategies reduced the effective reproduction number more than mass testing or self-isolation alone (Table 3). If only self-isolation of symptomatic cases was implemented in the model, it resulted in a mean reduction in transmission of 32%. The addition of household quarantine to self-isolation resulted in an overall mean reduction of 37%. In our simulations, self-isolation combined with manual contact tracing of all contacts reduced transmission by 61%; manual tracing of acquaintances only (i.e. contacts that had been met before) led to a 57% reduction in transmission. We estimated that self-isolation combined with app-based tracing with our baseline assumption of 53% coverage reduced transmission by around 44%, because both the primary case and contact would need to have the app to successfully quarantine an infected secondary case. Contact tracing measures also substantially reduced the probability that a primary symptomatic case would generate more than one secondary case (Table 3).

**Table 3:** Mean reduction in effective reproduction number under different control measures (i.e. the relative reduction from quarantining infectious individuals that would have gone undetected with no intervention). Results from 20,000 simulated setting-specific secondary transmission, assuming secondary attack rate of 20% among household contacts and 6% among other contacts. Results under the assumption of some workplace restrictions remaining in place are shown in Table 4. Estimates are shown to two significant figures. HH = household.

| **Scenario** | **Self-Isolation (SI)** | **Contact tracing** | **% non-HH contacts that are potentially traceable** | **% cases that have $R > 1$** | **$R_{\text{eff}}$** | **Mean reduction in $R_{\text{eff}}$** |
| --- | --- | --- | --- | --- | --- | --- |
| No control | No | No | – | 50% | 2.6 | 0% |
| Self-isolation (SI) | Yes | No | – | 38% | 1.7 | 32% |
| SI & HH quarantine | Yes | HH | – | 35% | 1.6 | 37% |
| SI, HH quarantine + work/school contact tracing | Yes | HH & work/school | 90% school, 79% work, 52% other | 28% | 1.2 | 52% |
| SI + manual CT of acquaintances | Yes | All | 90% school, 79% work, 52% other | 25% | 1.1 | 57% |
| SI + manual CT of acquaintances + limit to 4 daily ‘other’ contacts | Yes | All | 90% school, 79% work, 52% other | 21% | 0.91 | 64% |
| SI + manual contact tracing of all contacts | Yes | All | 100% | 22% | 1 | 61% |
| SI + app-based tracing | Yes | All | 53% | 31% | 1.4 | 44% |
| SI + app-based tracing + limit to 4 daily ‘other’ contacts | Yes | All | 53% | 27% | 1.2 | 53% |
| Mass weekly population testing | No | – | – | 50% | 2.5 | 2.20% |

We estimated that if some level of physical distancing were maintained, it could supplement the reduction in transmission from contact tracing. For example, if daily contacts in ‘other’ settings (i.e. outside the home, work and school) were limited to four people (the mean number made in our dataset), our model suggested that manual tracing of acquaintances only could lead to a 64% reduction in transmission, and app-based tracing a 53% reduction. We estimated that mass random testing of 5% of the population each week would reduce transmission by only 2%, because relatively few infections would be detected and many of those that were would have already spread infection to others.

We also considered the number of contacts that would be traced under different strategies. In a scenario where there were 20,000 new symptomatic cases per day, most contact tracing strategies would require over 200,000 contacts to be newly quarantined each day on average as a result (Table 4). If incidence was at a lower level of 5,000 new symptomatic cases per day, there would be a corresponding four-fold reduction in the number of daily contacts that needed to be quarantined. Although there was a similar reduction in transmission from manual tracing of all contacts and manual tracing of only acquaintances with a limit to four daily contacts in other settings (Table 3), the latter combination required fewer people to be quarantined each day (Table 4). We obtained similar results for the relative reductions in transmission and number of contact traced when we assumed a higher secondary attack rate within-household or among other contacts, which corresponded to baseline reproduction numbers of 2.6–3 (Table S1).

**Table 4:** Numbers of additional people quarantined per symptomatic case under different assumptions about new symptomatic cases per day. We assume quarantined contacts are independent. Estimates shown to two significant figures, with median and 90% prediction interval given for additional contacts quarantined per detected symptomatic case.

| Scenario | Number of people quarantined per detected case (median, 90% PI) | Mean newly quarantined per day (thousands) assuming 20k new symptomatic cases per day. | Mean newly quarantined per day (thousands) assuming 10k new symptomatic cases per day. | Mean newly quarantined per day (thousands) assuming 5k new symptomatic cases per day. |
| --- | --- | --- | --- | --- |
| SI & HH quarantine | 2 (0-7) | 43 | 22 | 11 |
| SI, HH quarantine + work/school contact tracing | 12 (0-100) | 500 | 250 | 130 |
| SI + manual CT of acquaintances | 21 (1-120) | 650 | 320 | 160 |
| SI + manual CT of acquaintances + limit to 4 daily 'other' contacts | 17 (1-110) | 580 | 290 | 140 |
| SI + manual contact tracing of all contacts | 28 (1-130) | 770 | 390 | 190 |
| SI + app-based tracing | 4 (0-57) | 270 | 140 | 68 |
| SI + app-based tracing + limit to 4 daily 'other' contacts | 4 (0-46) | 220 | 110 | 54 |

We found that the effectiveness of manual contact tracing strategies were highly dependent on how many contacts were successfully traced, with a high level of tracing required to ensure *R*_*eff*_<1 in our baseline scenario (Figure 2A). If contact tracing were combined with a maximum limit to daily contacts made in other settings (e.g. by restricting events), we found that this limit would have to be relatively small (i.e. fewer than 10–20 contacts) before a discernible effect could be seen on *R*_*eff*_. The limit would have to be very small (i.e. fewer than around 10 contacts) to ensure *R*_*eff*_<1 for app-based tracing, even if half of adults also had no work contacts because remote working was in place (Figure 2B). When app-based tracing is in place, we estimated that if only work contacts are restricted, a substantial proportion of the adult population would need to have zero work contacts to ensure *R*_*eff*_<1 (Figure 2C). Under our baseline assumptions, we estimated that app-based tracing would require a high level of coverage to ensure *R*_*eff*_<1 (Figure 2D), because both primary case and contacts would need the app; this is consistent with our finding that manual tracing would require a high proportion of contacts to be traced.

**Figure 2:**
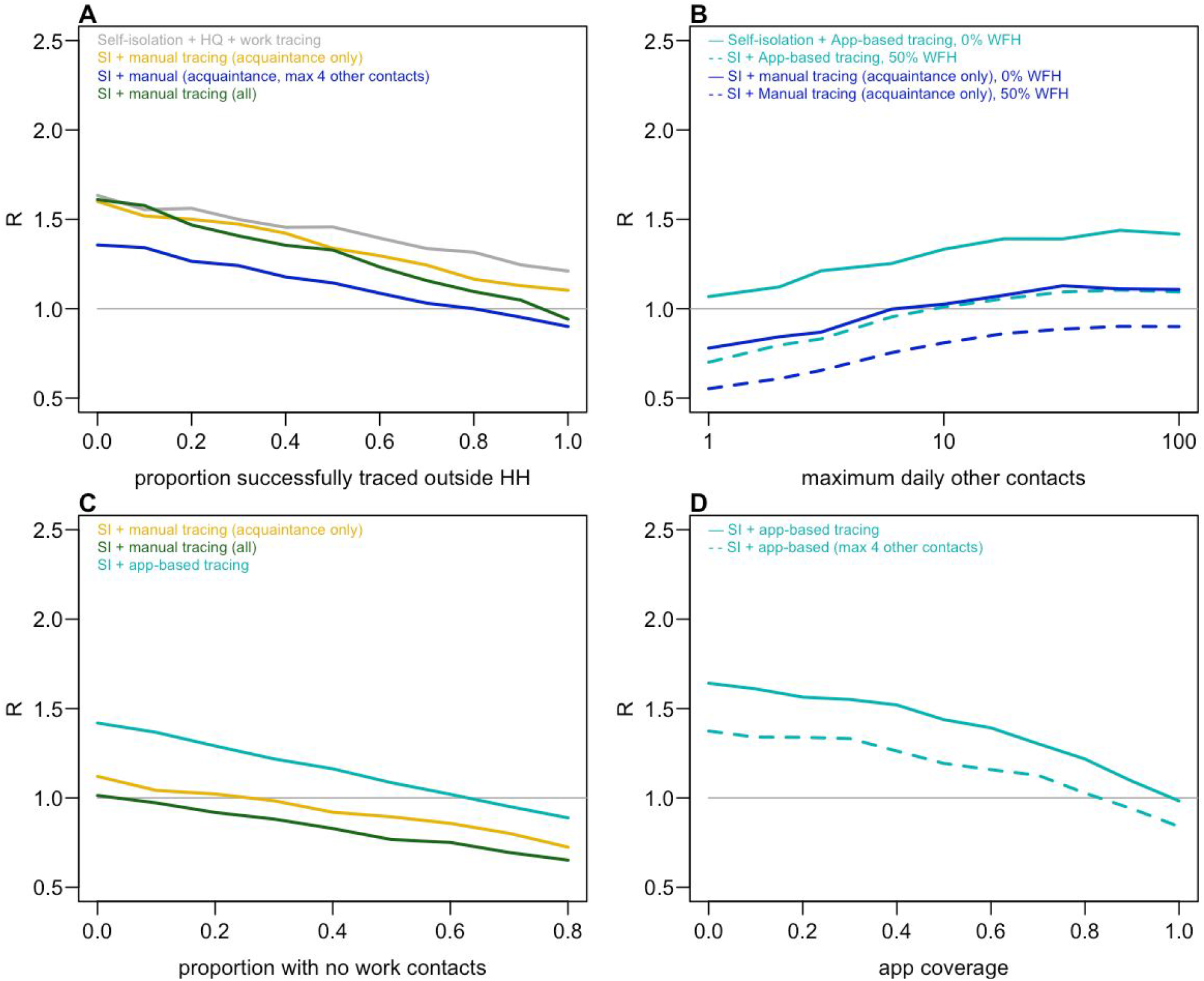
Impact of contact tracing effectiveness and physical distancing on reduction in reproduction number (baseline R=2.6). A) Reduction in R under different strategies for different proportions of work/school/other contacts that are successfully traced. B) Effect of the maximum limit on the number of daily contacts in other settings and control tracing strategies on R, either when adults are working as normal, or when 50% have no work contacts (WFH=50%). C) Effect of proportion of population with no work contacts. D) Effect of app-based tracing under different assumptions about app coverage. In all panels, other parameters are as in Table 2.

We also considered the impact of assumptions about the proportion of infections which are symptomatic and the relative contribution of asymptomatic individuals to transmission. We estimated that if a high proportion of cases were symptomatic, self-isolation and contact tracing measures would lead to a greater relative reduction in transmission (Figure S2A); this is mostly because more primary cases would be detected. Control measures were slightly less effective if the relative transmissibility of asymptomatic infections was higher (Figure S2B), because it would mean more undetectable transmission. However, because our baseline scenario assumed 60% of cases were symptomatic, the overall effect was less than it would be if the majority of cases were asymptomatic. We estimated that if there was no pre-symptomatic transmission, or individuals self-isolated rapidly (i.e. with 1.2 days on average rather than 2.6 days), self-isolation and household quarantine would lead to a larger reduction in transmission (Table S2); correspondingly, if we assumed cases took longer to self-isolate after becoming symptomatic (i.e. 3.6 days on average), these measures were less effective. However, the estimated overall reduction from self-isolation and manual contact tracing was similar across the three scenarios, because although more secondary infections occurred before isolation, a large proportion of them would be traced under our baseline model assumptions.

## Discussion

Using a model of setting-specific interactions, we estimated that strategies that combined isolation of symptomatic cases and tracing of their contacts reduced the effective reproduction number more than mass testing or self-isolation alone. The effectiveness of these isolation and tracing strategies was further enhanced when combined with physical distancing measures, such as a reduction in work contacts, or a limit to the number of contacts made outside of home, school or work settings. Several countries have achieved a prolonged suppression of SARS-CoV-2 transmission using a combination of case isolation, contact tracing and physical distancing. In Hong Kong, isolation of cases and tracing of contacts was combined with other physical distancing measures, which resulted in an estimated effective reproduction number near 1 throughout February and March 2020 (10). As well as early surveillance and containment measures (2), Singapore introduced additional ‘circuit breaker’ interventions to counter growing case numbers in April 2020 (23). In South Korea, testing and tracing has been combined with school closures and remote working (24,25).

In our analysis, we estimated that a large number of contacts would need to be traced and tested if incidence of symptomatic cases was high. This logistical constraint may influence how and when it is possible to transition from ensuring *R*_*eff*_<1 through extensive physical distancing measures to reducing transmission predominantly through targeted isolation and tracing-based measures. Our estimate of 20–30 median contacts being traced and tested per case in the manual tracing strategies we considered (Table 4) is reflected in the large scale testing being conducted per confirmed case in countries with a high estimated proportion of cases reported as of mid-April 2020, such as Australia (64 tests per case) and South Korea (52 tests per case) (26,27). This suggests any planning for ongoing control based on isolation and tracing should account for the likely need to conduct at least 30–50 additional tests for each case detected.

Our analysis has several limitations. We focused on individual-level transmission between a primary case and their contacts, rather than considering higher degree network effects. If contacts were clustered (i.e. know each other), it could reduce the number of contacts that need to be traced over multiple generations of transmission. We also assumed that contacts made within the home are the same people daily, but contacts outside home are made independently each day. Repeated contacts would also reduce the number that need to be traced. However, our estimates are consistent with the upper bound of numbers traced in empirical studies (Table 1), as well as analysis of UK social interactions that accounts for higher degree contacts (14). Because our data was not stratified beyond the four contact settings we considered (home, work, school, other), we could not consider further specific settings, e.g. mass gatherings. However, our finding that gatherings in other settings needed to be restricted to relatively small sizes before there was a noticeable impact on transmission is consistent with findings that groups between 10–50 people have a larger impact on SARS-CoV-2 dynamics than groups of more than 50 (28).

Our baseline assumptions were plausible but optimistic. In particular, we assume a delay of symptom onset to isolation of 2.6 days in the baseline scenario, and quarantine that was sufficiently fast to prevent any onwards transmission among successfully traced contacts, with 90% assumed to adhere to quarantine. Based on viral shedding dynamics, this would imply tracing and quarantine within around 2–3 days of exposure (6). We also assumed that routine self-isolation would not increase household transmission. However, our conclusions about onwards transmission in the different control tracing scenarios would not be affected if some limited household transmission did occur, because in these scenarios we assumed that household quarantine would be in place too. We also simulate contact patterns at random for each individual in our population, whereas in an outbreak, there is likely to be a correlation between degree and infection risk; individuals with multiple contacts may be more likely to acquire infection as well as transmit to others (29). If this were the case, and we assume the same secondary attack rates, the overall reduction may be lower than we have estimated; however, to keep the baseline reproduction number consistent, this correlation would have to be offset by a lower SAR among contacts.

Our results highlight the challenges involved in controlling SARS-CoV-2. Consistent with previous modelling studies (7,14) and observed early global outbreak dynamics, our analysis suggests that, depending on the overall effectiveness of testing, tracing, isolation and quarantine, a combination of self-isolation, contact tracing and physical distancing may be required to ensure *R*_*eff*_<1. Further, in a scenario where incidence is high, a considerable number of individuals may need to be quarantined to achieve control using strategies that involve contact tracing.

## Data Availability

Data and code are fully available.

https://github.com/adamkucharski/2020-cov-tracing

## Supplementary Information

**Figure S1:**
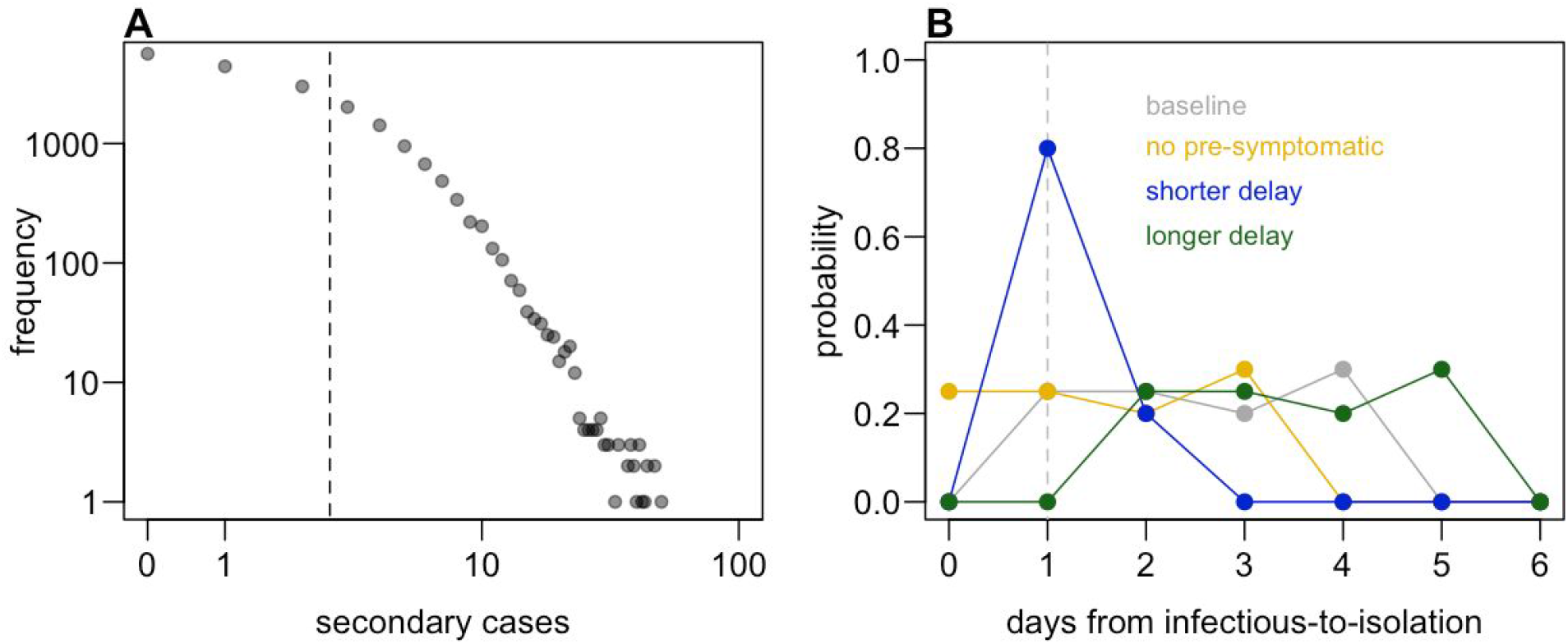
Model assumptions about transmission and infectiousness. A) Individual-level distribution of secondary transmission in baseline scenario. Dashed line shows mean (i.e. R_eff_). B) Distribution scenarios for delay from infectious-to-isolation. In scenarios with pre-symptomatic transmission, we assume this period lasts one day; dashed line shows time of onset of symptoms in these scenarios.

**Figure S2:**
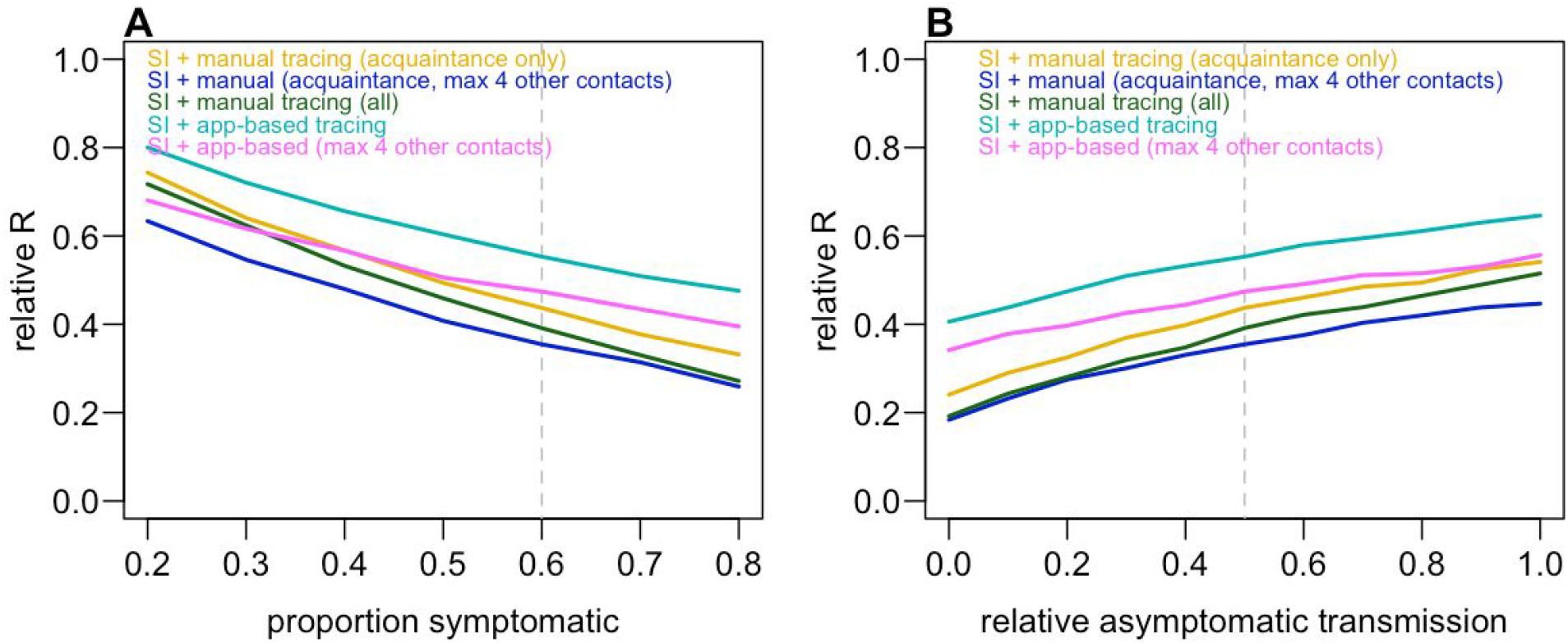
Impact of proportion of the population who are symptomatic and relative transmission from asymptomatic individuals on reduction in transmission. A) Relative reduction in the reproduction number (i.e. ratio between baseline R and R under control measures) when different proportions of the population are symptomatic. B) Relative transmission reduction when asymptomatic individuals have different relative transmission risks compared to symptomatic individuals. Dashed lines show baseline assumption.

**Table S1:** Reduction in transmission and number of contacts quarantined per symptomatic case under different assumptions about secondary attack rate (SAR) among contacts made within and outside households. Median and 90% prediction interval shown for contacts quarantined. HH SAR=20% and other contact SAR=7% corresponded to baseline *R*_*eff*_=3; HH SAR=40% and other contact SAR=5% corresponded to baseline *R*_*eff*_=2.6.

| Scenario | Baseline assumptions |  | HH SAR=20%, other contact SAR=7% |  | HH SAR=40%, other contact SAR=5% |  |
| --- | --- | --- | --- | --- | --- | --- |
|  | Reduction | Quar. | Reduction | Quar. | Reduction | Quar. |
| Self-isolation (SI) | 32% | – | 33% | – | 33% | – |
| SI & HH quarantine | 37% | 2 (0-7) | 38% | 2 (0-7) | 42% | 2 (0-7) |
| SI, HH quarantine + work/school contact tracing | 52% | 12 (0-100) | 51% | 12 (0-100) | 54% | 13 (0-99) |
| SI + manual CT of acquaintances | 57% | 21 (1-120) | 57% | 22 (1-120) | 58% | 22 (1-120) |
| SI + manual CT of acquaintances + limit to 4 daily ‘other’ contacts | 64% | 17 (1-110) | 64% | 18 (1-100) | 64% | 18 (1-110) |
| SI + manual contact tracing of all contacts | 61% | 28 (1-130) | 61% | 28 (1-130) | 61% | 28 (1-130) |
| SI + app-based tracing | 44% | 4 (0-57) | 45% | 5 (0-59) | 48% | 5 (0-57) |
| SI + app-based tracing + limit to 4 daily ‘other’ contacts | 53% | 4 (0-46) | 53% | 5 (0-46) | 56% | 4 (0-46) |

**Table S2:** Reduction in transmission and number of contacts quarantined per symptomatic case under different assumptions about pre-symptomatic period and delay to self-isolation. Assumptions about the distributions of delays shown in Figure S1. Median and 90% prediction interval shown for contacts quarantined.

| Scenario | Baseline assumptions |  | No pre-symptomatic transmission |  | Shorter delay to self-isolation |  | Longer delay to self-isolation |  |
| --- | --- | --- | --- | --- | --- | --- | --- | --- |
|  | Reduction | Quar. | Reduction | Quar. | Reduction | Quar. | Reduction | Quar. |
| Self-isolation (SI) | 32% | – | 46% | – | 51% | – | 20% | – |
| SI & HH quarantine | 37% | 2 (0-7) | 50% | 2 (0-7) | 54% | 2 (0-7) | 26% | 2 (0-7) |
| SI, HH quarantine + work/school contact tracing | 52% | 12 (0-100) | 57% | 13 (0-100) | 60% | 12 (0-100) | 46% | 13 (0-100) |
| SI + manual CT of acquaintances | 57% | 21 (1-120) | 62% | 22 (1-120) | 62% | 21 (1-110) | 53% | 22 (1-110) |
| SI + manual CT of acquaintances + limit to 4 daily ‘other’ contacts | 64% | 17 (1-110) | 67% | 18 (1-100) | 69% | 18 (1-110) | 61% | 17 (1-100) |
| SI + manual contact tracing of all contacts | 61% | 28 (1-130) | 64% | 28 (1-130) | 64% | 28 (1-130) | 59% | 28 (1-130) |
| SI + app-based tracing | 44% | 4 (0-57) | 53% | 4 (0-57) | 57% | 4 (0-58) | 36% | 4 (0-59) |
| SI + app-based tracing + limit to 4 daily ‘other’ contacts | 53% | 4 (0-46) | 61% | 5 (0-45) | 63% | 5 (0-46) | 46% | 5 (0-45) |

## Notes

### Competing Interest Statement

The authors have declared no competing interest.

### Funding Statement

The work was supported by Wellcome Trust (206250/Z/17/Z), European Commission (101003688) and EPSRC (EP/N509620/1).

